# Visual Transformer and Deep CNN Prediction of High-risk COVID-19 Infected Patients using Fusion of CT Images and Clinical Data

**DOI:** 10.1101/2022.07.26.22278084

**Authors:** Sara Saberi Moghadam Tehrani, Maral Zarvani, Paria Amiri, Reza Azmi, Zahra Ghods, Narges Nourozi, Masoomeh Raoufi, Seyed Amir Ahmad Safavi-Naini, Amirali Soheili, Sara Abolghasemi, Mohammad Gharib, Hamid Abbasi

**Affiliations:** Faculty of Engineering, Alzahra University, Tehran, Iran; Pooyandegan Rah Saadat Company, Tehran, Iran; Department of Radiology, School of Medicine, Imam Hossein Hospital, Shahid Beheshti, University of Medical Sciences, Tehran, Iran; Research Institute for Gastroenterology and Liver Diseases, Shahid Beheshti University of Medical Sciences, Tehran, Iran; School of Medicine, Shahid Beheshti University of Medical Sciences, Tehran, Iran; Infectious Diseases and Tropical Medicine Research Center, Shahid Beheshti University of Medial Sciences, Tehran, Iran; Auckland City Hospital, Auckland, 1010, New Zealand; Auckland Bioengineering Institute, University of Auckland, Auckland, 1010, New Zealand

**Keywords:** Deep Learning, Visual Transformer, Predictive models, convolutional neural network (CNN), Covid-19 detection, CT scan, clinical data, data fusion

## Abstract

Despite the globally reducing hospitalization rates and the much lower risks of Covid-19 mortality, accurate diagnosis of the infection stage and prediction of outcomes are clinically of interest. Advanced current technology can facilitate automating the process and help identifying those who are at higher risks of developing severe illness. Deep-learning schemes including Visual Transformer and Convolutional Neural Networks (CNNs), in particular, are shown to be powerful tools for predicting clinical outcomes when fed with either CT scan images or clinical data of patients.

This paper demonstrates how a novel 3D data fusion approach through concatenating CT scan images with patients’ clinical data can remarkably improve the performance of Visual Transformer and CNN models in predicting Covid-19 infection outcomes. Here, we explore and represent comprehensive research on the efficiency of Video Swin Transformers and a number of CNN models fed with fusion datasets and CT scans only *vs* a set of conventional classifiers fed with patients’ clinical data only. A relatively large clinical dataset from 380 Covid-19 diagnosed patients was used to train/test the models. Results show that the 3D Video Swin Transformers fed with the fusion datasets of 64 sectional CT scans+67 (or 30 selected) clinical labels outperformed all other approaches for predicting outcomes in Covid-19-infected patients amongst all techniques (i.e., TPR=0.95, FPR=0.40, F0.5 score=0.82, AUC=0.77, Kappa=0.6). Results indicate possibilities of predicting the severity of outcome using patients’ CT images and clinical data collected at the time of admission to hospital.

## 1. Introduction

In the late 2019, Covid-19 pandemic was initially reported to rapidly infect residents of Wuhan city in China [1]. This previously unknown virus was then labelled as SARS-CoV2 by the International Committee on Taxonomy of Viruses (ICTV) and categorized under the family of corona viruses [2]. The infection caused by the Covid-19 was reported to be very similar to the disease due to the infection by SARS virus and could lead to severe respiratory syndromes and death [3, 4]. The fast and large increase in the number of infected individuals before vaccine roll-outs had resulted in a large increase in the number of referrals with critical conditions and admittance to the hospitals and clinics, imposing a burden on the healthcare sector, globally. This important factor could potentially result in an increase in critical human error that could lower the diagnosis accuracy, subsequently. Recent analytical enhancements could assist in finding practical solutions to the urgent need for developing automated diagnosis platforms that can provide prognostic information about the evolution of infection in patients. Clinical observations confirm a large variety of symptoms for the infected individuals, where the milder initial symptoms could rapidly develop to critical situations. This itself could limit the clinical assessments or in more severe cases can eliminate the chances of treatment [5]. Therefore, clinical monitoring of patients and accurate prediction of infection development during this period and/or even before their initial referrals can play an important role in saving lives [6]. Research suggest that the quality of patients’ chest Computerized Tomography (CT) scans are interpretably linked to other observations from patients including their clinical examinations, laboratory tests, vital signals, patient history, and potential background illnesses [7]. Therefore, it is hypothesized that a proper combination of these data could be used for automatic prediction of both the severity and the developmental stage of the infection, more accurately [8].

Various applications of multi-modal data fusion techniques on Covid datasets have been addressed in the literature. Studies suggest that chest X-ray images and lung CT scans can be fed into deep-learning-based models for diagnosis and classification of Covid-19-related conditions [9-12]. Access to larger clinical datasets is currently a major challenge in the implementation of these techniques. Thus, various research have considered data augmentation techniques to cover these drawbacks [13-15]. Attempts show that predictive models fed with patients’ clinical data, demographic/historical conditions and disorders, as well as laboratory tests can be used to predict outcomes [15-17]. Literature indicates possibilities of developing high-performance algorithms to accurately predict the severity of infection and further diagnose healthy individuals from tested-positive cases. Successful algorithms have used combinational approaches through fusioning clinical observations data, CT images, vital signals, and background/historical conditions [8, 17-20]. These studies have initially combined features extracted from CT images with features from the patients’ clinical data and fed the outputs into deep-net classifiers. For instance, studies show that the extracted features from the images can be combined with other available features/data (e.g., clinical observations/measures) to create a more robust and consistent dataset that can provide detailed information for the deep-net to predict the severity of infection in the high- and low-risk patients [8, 14, 21].

In this work, we use data fusion of lung CT scan images and clinical data from a total of 380 Iranian Covid-19-positive patients to develop deep-learning-based models to predict risk of mortality and outcomes in the high-vs low-risk Covid-19 infected individuals. An overall schematic of the proposed approaches in this work is shown in Figure 1. The article contributes to the field through:

**Figure 1.**
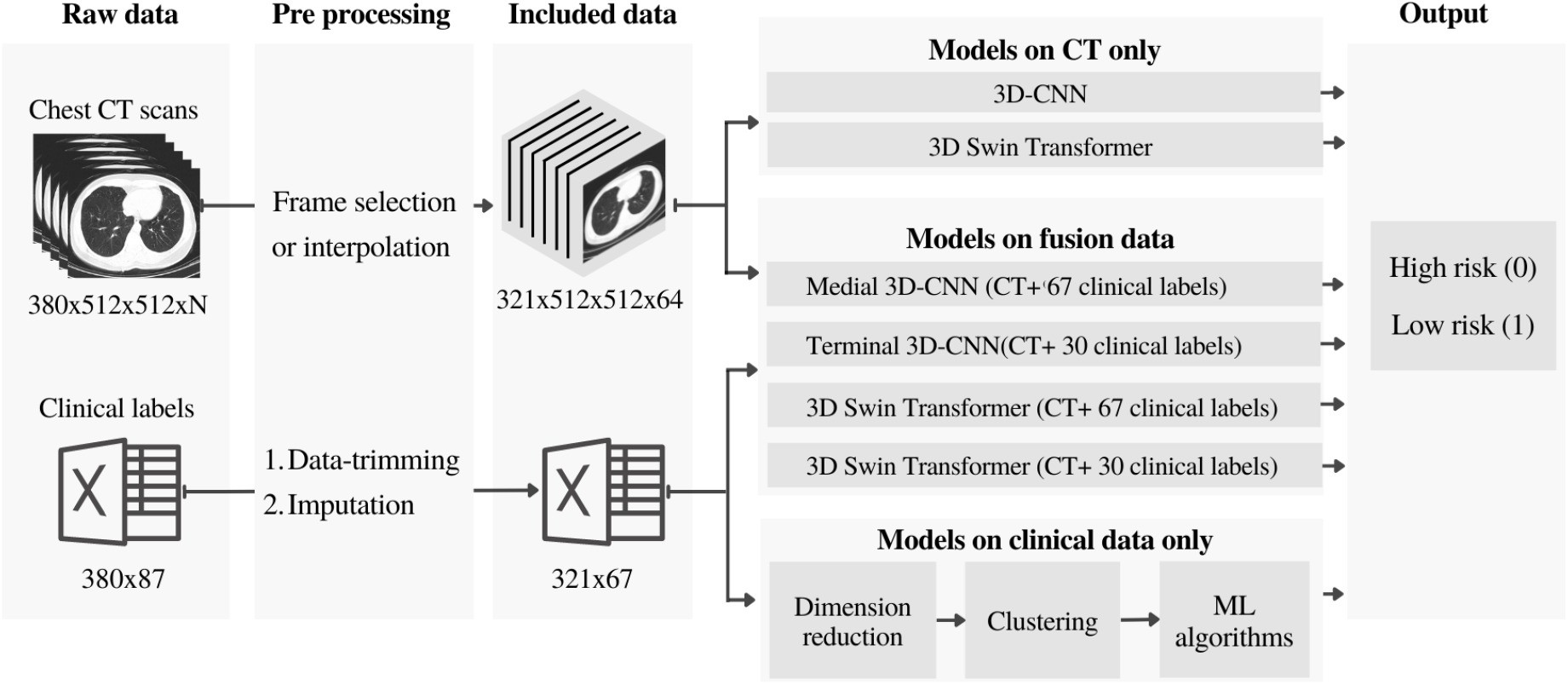
The flow-chart schematic of the proposed predictive machine-learning approaches for the classification of high- and low-risk Covid-19 infected patients. “N” denotes the number of total CT scan slices from each patient.

1. Developing Visual Transformer and 3D Convolutional Neural Network (CNN) predictive models fed with a series of fusion datasets from patients’ CT images and their clinical data. This includes introducing a novel heuristic concatenation approach, for integrating CT scan images with clinical data, which is inferred to have assisted with inter-network feature aggregations in the Transformer models.
2. Developing Visual Transformer and CNN-based predictive models fed with CT scan images only, and assessing the capabilities of genetic algorithm (GA) for hyper-parameter tuning of the 3D-CNN models fed with the fusion data and CT scan images.
3. Evaluating a series of conventional classifiers for predicting outcomes using patients’ clinical data only, and investigating strategies to select a set of proper clinical labels from the pool of clinical data for the classification of imbalance data. The paper further discusses imputation techniques to deal with missing values in the dataset.

## 2. Related work

### 2.1. Clinical data-based detection

Here, only patients’ clinical data, including patients’ history and their lab test results, are used to develop predictive models. Yue et al. have demonstrated that the use of clinical data and patients’ condition assessments at the time of admission can help to predict chances of mortality at around 20 days [14]. They have achieved promising results by integrating predictive models including logistic regression (LR), support vector machine (SVM), gradient boosted decision tree (GBDT), and neural networks (NN) to predict the mortality risk (AUC: 0.924-0.976) [14]. Dhruv et al., have also shown that patients’ clinical data, blood panel profiles, and socio-demographic data can be fed into conventional classification algorithms such as Extra Tree, gradient boosting, and random forest for predicting the severity of Covid-19 [15]. Similar works show that clinical parameters in the blood samples can be infused into a combined statistical analysis and deep-learning model to predict severity of Covid-19 symptoms and classify healthy individuals from tested-positive cases [17].

### 2.2. Image-based detection

In this approach, only chest X-ray or CT scan images are used for classification of Covid-19 infected patients. Purohit et al. have proposed an image-based Covid-19 classification algorithm and demonstrated that, among various image sharpening techniques, utilization of certain sharpening filters such as canny, sobel, texton gradient and their combinations can help to increase training accuracy in multi-image augmented CNN [13]. Research shows that deep neural networks are able to automatically diagnose Covid-19 infection in partial X-ray images of the lungs [22], or through fusioning deep features of CT images [23-25]. Our team has also previously shown that chest X-ray images can be fed into CNNs for Covid detection [26].

Visual Transformer (ViT) networks, along with the CNN models, have recently shown remarkable capability in resulting higher performances in various applications, such as image classification, object detection, and semantic segmentation. Recent works show that ViT and in particular Video Swin Transformers can competitively achieve better accuracies, compared to the CNN-based methods, for the classification and identification of Covid-19 infected patients using chest CT scans [27] and X-ray images [28]. Research shows that the feature maps extracted from the CT scan images in the output of a ResNet model can be used as inputs to a transformer model for the identification of Covid patients (∼1934 images, >1000 patients, recall accuracy 0.93) [29]. Transfer-learning in Visual Transformer models, fed with either CT images or their combinations with chest X-ray images, shows diagnostic possibilities of Covid-19 patients and localization of the infected regions in the lungs [28, 30]. A recent work has shown that a combination of parallelly extracted features from CT scans through simultaneous application of Visual Transformers and CNN can help to accurately classify Covid-19 patients [31]. Fan et al. have reported a high recall performance of 0.96 using 194,922 images from 3745 patients which suggests strong capabilities of combinational approaches [31].

### 2.3. Fusion-based detection

This approach mainly aims to fuse patients’ clinical data with any other possible information, such as chest X-ray and/or CT images, to use as the inputs for predictive models. Using a relatively large CT image dataset from multiple institutions across three continents, Gong et al. have developed a deep-learning-based image processing approach for diagnosis of Covid-19 lung infection [18]. In their technique, a deep-learning model initially segments lung infected regions by extracting total opacity ratio and consolidation ratio parameters from CT images and then combines the outputs with clinical and laboratory data for prognosis purposes using a generalized linear model technique (reported AUC range: 0.85–0.93) [18]. Other studies have proposed robust 3D CNN predictive models fed with combined data from segmented CT images and patients’ clinical data to predict whether a Covid-19-infected-individual belongs to the low- or high-risk group [8, 19, 20]. These studies have shown that their proposed approaches are independent of demographic information such as age and sex, and other conditions such as chronic diseases. Meng et al., have demonstrated that 3D-CNNs can perform much better when simultaneously fed with patients’ segmented CT scans and clinical data compared to the singular use of clinical data or CT images in CNN-based or logistic regression models [19]. Ho et al., have compared performances of three 3D CNNs where each was trained on the 1) raw CT images, 2) segmented CT images, and 3) on the long lesion segmented data. They reported higher performance from the last approach amongst all [20].

A recent study has initially trained a speech identification model for Covid diagnosis using Long Short-Term Memory Networks (LSTM) that uses the acoustic aspects of patient’s voice, their breathing data, coughing patterns, and talking [32]. The patients’ chest X-ray images are also fed into general deep-net models, including a VGG16, a VGG19, a Densnet201, a ResNet50, a Inceptionv3, a InceptionResNetV2, and a Xception for Covid identification. Images and audio features were then combined and used as inputs to a hybrid model to identify non-Covid or Covid-positive patients. They have reported a lower accuracy for their hybrid model compared to their speech-based or X-ray image-based models [32].

## 3. Methods and computational approach

### 3.1. Dataset

The dataset used in this research includes both lung CT scan images and their clinical data from a total of 380 Covid-19 infected patients. Patients were diagnosed by clinicians according to the Iran’s National Health guidelines [33] through clinical assessments of their symptoms and lung CT images. The patients were hospitalized in the emergency unit at Imam Hussain Hospital, Tehran, Iran, between 22nd Feb 2020 to 22nd March 2020. All ethics of the current research have been approved by the Shahid Beheshti University’s ethics committee (Ref: IR.SBMU.RETECH.REC.1399.003). All patients have signed and submitted their consent to participate in the research and their data privacy has been fully considered [34]. Examples of the lung CT scans at different slice locations from a high- and a low-risk patient are shown in Figure 2A, 2B and 2C, 2D, respectively.

**Figure 2.**
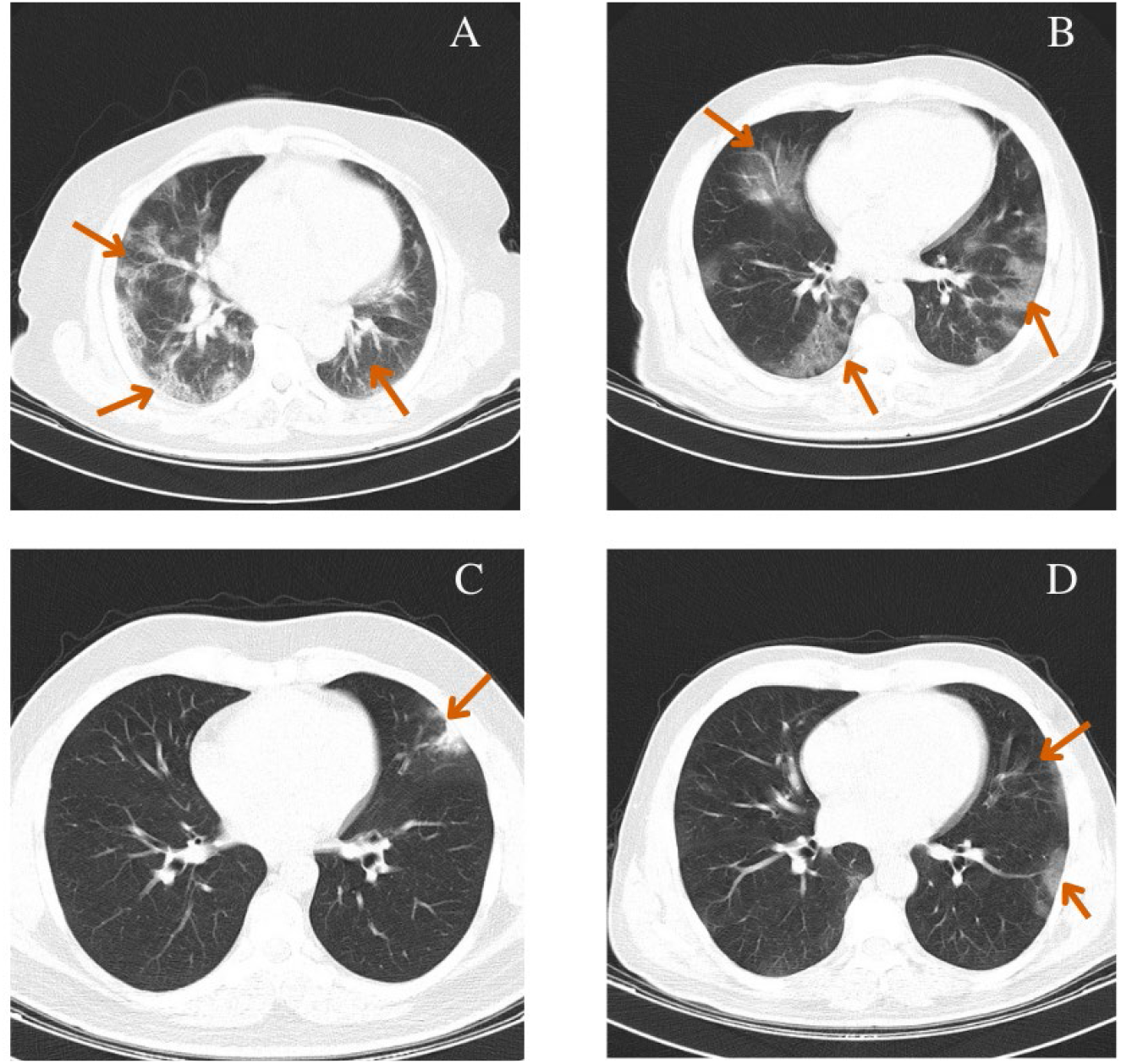
Examples of the lung CT scans from the sequence of slices in the high- (A, B) and low-risk (C, D) Covid-19 infected patients. Arrows indicate infected regions.

From the total number of studied patients, 318 individuals have recovered from the illness while 62 have died. Since our top goal in this research was to correctly predict the severity of outcomes (mortality risk) using data collected at or around the time of initial referral, we categorized died patients (including ICU-hospitalized deaths) in the high-risk group (class 0) and labeled the recovered individuals as the low-risk class (class 1).

Our image datasets included a series of lung CT scans ranging between 50 to 70 images/slices depending on the length of the patient’s lung. The clinical data used in this research were used as sets of numerical data collected for all patients, including demographic data, exposure history, background illness or comorbid diseases, symptoms, presenting vital signs, and laboratory tests data. A full list of these parameters as well as their mean and standard deviation (mean±std) are tabulated in Table A.1 of Appendix A.

### 3.2. Data pre-processing

An optimal data pre-processing is a critical initial step, prior to the initiation of training process, with possible boosting impacts on the overall performance of a model. A variety of pre-processing strategies can be chosen based on the type of data and/or algorithms used. In the following, we detail our pre-processing approaches for the numerical datasets and CT images.

#### 3.2.1 Pre-processing of clinical data

Dealing with clinical data are often associated with certain challenges. For example, finding appropriate values for the missing data requires strategic imputations or conversion of qualitative measurements to numerical formats. Data often holds high dimensionality and strategies such as dimension reduction (data/feature/label selection) and feature extraction can help to represent data in a simpler format. Initially, we used one-hot encoding [35] approach to convert qualitative data such as gender, etc., into numerical representations. In the following, we explain our strategies for clinical data trimming and preparation for analysis.

##### 3.2.1.1 Clinical data trimming

Initially, we considered a thresholding criterion to remove patients from the original clinical dataset, whom at least 55% of their clinical data was missing (59 patients were removed). To improve data clarity and robustness, we further trimmed the dataset by removing clinical labels with at least 60% missing values for the entire dataset (20 labels were removed). These threshold values were chosen based on manual assessments and observations. From the remaining 321 patients (total of 380 patients), those who died were categorized as “high-risk” (n=57, class 0) and the remaining participants were labeled as “low-risk” (n=264, class 1).

##### 3.2.1.2 Imputation of missing values

The intensive work-load of clinical staff or other emergency situations/reasons may lead to missing values in patients’ data recordings. Therefore, clinical datasets are often imputed to cover the missing information on the sheets. In the machine-learning field, it is also challenging to work with the initial format of the imputed datasets. Therefore, it is inevitable to use proper algorithms to fill in for the missing data or remove some [36]. Research shows that statistical approaches can be used to estimate the missing data using statistical parameters, such as mean, median, etc., from the entire dataset. Also, other machine-learning techniques such as linear regression or k-nearest neighbor (KNN) can be used to estimate the missing values [37-41]. Here, we used KNN algorithm (k=5) to provide estimations for the missing values in the dataset.

##### 3.2.1.3 Dimension reduction

Generally, dimension reduction is performed via feature/label selection or feature extraction operations. Feature/label selection approaches are mainly concerned with distinguishing the most dominant features/labels while feature extraction strategies are employed to transfer data values into a new domain and sometimes define novel features based on the original ones. In this research, we investigated the impact of both approaches on the feature-sets and assessed outcomes for each, both visually and by implementing a set of conventional classifiers explained in the following. The final extracted features as well as the selected clinical labels from these attempts were later used in the training process. In this study, we often refer to the selected clinical data as “clinical labels”.

<u>Feature extraction</u>: here, we extracted features from the clinical data by utilizing a commonly used dimension reduction technique, namely called principal component analysis (PCA) [42, 43]. PCA is an unsupervised and linear technique that uses eigen-vectors and eigen-values from a matrix of features to project lower dimensions from higher feature dimensions in the original matrix [44]. In the current study, an optimal number of required components in the PCA was found by using various numbers of extracted features. The output datasets from PCA were then fed into seven conventional classifiers including, SVM [38], MLP [46], KNN [47], random forest [48], gradient boosting [49], Gaussian naïve bayes [50], and XGBoost [51] to assess which number of feature-sets could lead to an optimal performance. This was accordingly found to be associated with a set of 25 components.

<u>Feature/label selection</u>: here we assessed the capabilities of two different approaches, namely “SelectKBest” [45, 46] and decision tree-based ensemble learning algorithms [47] to select a set of clinical labels from the pool of original clinical data. The SelectKBest algorithm uses statistical measures to score input features based on their relation to outputs and chooses the most effective features, accordingly. We used an ExtraTree classifier [48] for the decision tree-based ensemble learning approach where the algorithm randomly selects subsets of features to create the associated decision trees and evaluates minimal mathematical measures of each feature (typically the Gini Index [49]), while making the forest. Finally, all the extracted features are sorted in a descending order based on their measured Gini Index and user can choose to work with an arbitrary top *k* number of dominant features from the list. An optimal number of clinical labels was found by assessing the performance optimality of the aforementioned seven conventional classifiers across a set of various numbers for the SelectKBest algorithm and ExtraTree classifier. A set of 13 selected clinical labels from the SelectKBest and a set of 30 selected clinical labels from the ExtraTree classifier were found to result in better performances compared with other combination sets (see Appendix B).

We further visually assessed the selected features using an unsupervised non-linear technique based on manifold learning, called t-distributed stochastic neighbor embedding (t-SNE) [50]. The t-SNE is conventionally used for data visualization of large dimension datasets. t-SNE aims to find an optimized value for its cost function by measuring embedded similarities within the dataset at both higher- and lower-dimensions representations. In the t-SNE approach, a more visually separable data represents less complexity. Here, the t-SNE was applied to the 1) main dataset including all 67 clinical labels (with no dimension reduction), 2) a dataset including 13 selected clinical labels from SelectKBest, 2) a dataset including 30 selected clinical labels from ExtraTree classifier, and 4) a dataset including 25 extracted features from the PCA. Outputs of the t-SNE were then scatter plotted to visualize the complexity within each dataset (see results section for the plots). Finally, we chose to carry on with the set of 30 selected clinical labels from the ExtraTree classifier approach which were found to lead to better classification results and represented less visual complexity in the t-SNE approach.

#### 3.2.2 Pre-processing of CT images

CT scan images of lung consist of a sequence of video frames at various sections (slices) along the patient’s lung, where the number of frames varies in individuals according to their length of lung or device settings. These images can be used as the inputs for predictive/classification models where a certain number of input channels, that are compatible with the number of slices, must be used in the network’s architecture. Since the number of CT video frames varies across patients, an appropriate slice selection approach should be used to shape a uniform volumetric 3D input size for consistency across all models [51]. Various slice selection strategies consider manual selection of frames from the beginning, middle and end of a video set. The major problem with such approaches is that they neglect information connectivity across slices which can lead to loosing localized information and provide a false representation for the entire video set. On the other hand, there are strategies that initially select a fixed number of frames from the entire video, and then interpolate data to generate a desired set of frames that provides a more accurate representation of the whole video set [51] compared to the manual approach.

Here, we chose to work with Spline Interpolated Zoom (SIZ) frame selection technique. An arbitrary number of frames (N) is initially selected in this technique to construct a volumetrically uniform image-set that consists of a fixed number of CT slices for all individuals [51]. Then, depending on whether the patient’s video set contained higher or lower number of slices compared to the N, the sequence of slices was evenly sampled using a spacing factor or interpolated to construct the missing slices, respectively. Here, the original size of the gray scale CT images was provided as 512×512×1, and we chose to work with N=64 that represents the average number of frames in the videos from all patients. Therefore, the volumetrically uniform 3D inputs of the CNNs were re-shaped to the size of 512×512×64 for all patients.

#### 3.2.3 Data Fusion

Here, we combined the clinical data/measures with the 3D videos of the CT scan images from section 3.2.2 to shape more detailed fusion datasets for each patient. We initially expanded the dimension of clinical data through creating an empty 2D matrix with dimensions identical to the size of 2D video frames (i.e., 512×512). This 2D matrix was then replicated N times, where N is equal to the number of clinical labels. All data arrays of each 2D matrix were then filled with the value of the associated clinical label/measure. This 3D matrix of clinical data (512×512xN frames) was then added to the CT video (512×512×64 frames) to form a 3D fusion dataset of size 512×512x(64+N) frames. This dataset was then used as inputs to the model described in section 3.3.3.2, once with N=30 (suggested from 3.2.1.3) and with N=67.

### 3.3. Model Training

The training process for each of the four previously outlined models, in the introduction section and Figure 1, are described in the following:

#### 3.3.1 Approach #1: classification using clinical data only

Classification of datasets with imbalanced classes is associated with challenges and complexities which requires careful considerations. Data clustering is one of the useful approaches to handle such complexities and create more balanced datasets [52]. Here, we considered the following steps to create balanced datasets for training. Data were initially split into train and test sets (80% training, 20% test). The original ratio between class 1 and 0 in the clinical dataset is nearly 5 (imbalanced data), hence we used Gaussian mixture clustering algorithm [53] to divide the low-risk class in the training sets (n=264, class 1) into five different clusters. Each of these clusters were then combined with data from the high-risk class (n=57, class 0). This approach helped to create 5 separate balanced datasets which were then fed into seven different conventional classification algorithms, namely the SVM, MLP, KNN, random forest, gradient boosting, Gaussian naïve bayes, and XGBoost for classification. Each classification algorithm was accordingly trained and tested on the five balanced train/test datasets resulting in five classification measures for classifier (e.g., 5x trained/tested random forests). A voting approach was then applied to the outputs of these five blocks to determine the winning class. The class (e.g., 0 or 1) with a larger number of votes (i.e., 3, 4, 5) from all blocks were chosen as the winning class.

#### 3.3.2 Approach #2: training on CT images only

##### 3.3.2.1 3D-CNN CT model

Since the CT scan images are sequences of frames taken at different slices, therefore, they can be technically considered as 3D video data. Therefore, here we designed and trained a 3D-CNN with 3 convolutional layers on the training datasets. Here, inputs of the 3D-CNN classifiers are matrices of 512×512×64 dimension from the pre-processing stage, where 64 is the number of CT scan frames (slices) for each patient. We further used Genetic Algorithm (GA) to automatically find and assign optimal values for the CNNs’ hyperparameters [54]. The specified hyperparameters included the number of layers, number of neurons in each layer, learning-rate, optimization function, dropout size, and kernel size. The population size and the number of generation were set to 10 and 5, respectively. We also used Roulette wheel algorithm for parent selection followed by crossover mechanism. The GA were trained over 20 epochs and fitness values with lower FPRs were chosen, accordingly. The selected hyperparameters for the 3D-CNN-based models in this work are shown in Table C.1 in Appendix C.

##### 3.3.2.2 3D Swin Transformer CT model

Visual Transformers (ViT) are classes of deep neural networks that have been initially used for natural language processing (NLP) and sought as improved alternatives to other classes of deep-nets (i.e., CNNs) with competitive performances for multi-modal inputs [55]. An input image to a ViT is initially shaped as a set of image-patches (equivalent to the set of words in NLP) which is then embedded with the localized information of the image to form inputs to an encoder network within the Transformer. The encoder unit consists of a multi-head self-attention layer [56], which highly improves features learning such as long-range dependencies and aggregation of global information [57]. The multi-head self-attention layer therefore aggregates spatial locations’ information where global and local information are combined, accordingly. This operation is expected to help with inter-network feature extractions and lead to better outcomes compared to the CNN networks, where the receptive field sizes are fixed [58]. In CNNs, this can be equivalently achieved by increasing convolutional kernels which largely increases the computations. While ViT generally require 4 times lower computational facilities, they can extraordinary outperform the ordinary CNNs if trained on satisfactory data. Transformers generally require larger datasets for training, where transfer-learning and self-supervised techniques could greatly help to largely overcome such challenges. On the other hand, high-resolution input images can increase the computational burden and lower the computational speed, subsequently. To overcome this challenge, Swin Transfomer models have been introduced to deal with higher resolution data in computer vision applications [59]. Video Swin Transformer (VST) models have been further introduced to work with 3D datasets such as videos [60], where the application of transfer learning and pre-trained models have been helpful. In this work, we normalized and augmented the CT scan images of each patient from section 3.2.2 to form 3D inputs for a 3D Swin Transformer. With the aid of transfer-learning, we used a pre-trained model, Kinetics-400 [60], to set our model’s initial weights. We trained the 3D Swin Transformer over 50 epochs using an Adam optimizer with a 0.02 learning rate and 0.02 weight decay.

#### 3.3.3 Approach #3: training on fusion data

##### 3.3.3.1 3D-CNN models on fusion data

Here, we considered a terminal data fusion (on CTs+30 clinical labels) and a medial data fusion approach (on CTs+67 clinical labels) to combine the data. In the first approach, the terminal 3D-CNN, CT scans are initially fed into the 3D-CNN to extract their features-vector. The output features-vector were then terminally combined with the numerical data from dimension-reduction stage including 30 selected clinical labels for each patient to shape the final features-vector. The final features-vectors along with the labels were fed into the Naïve Bayes network [61] for training/classification. The schematic of this approach is shown in Figure 3.

**Figure 3.**
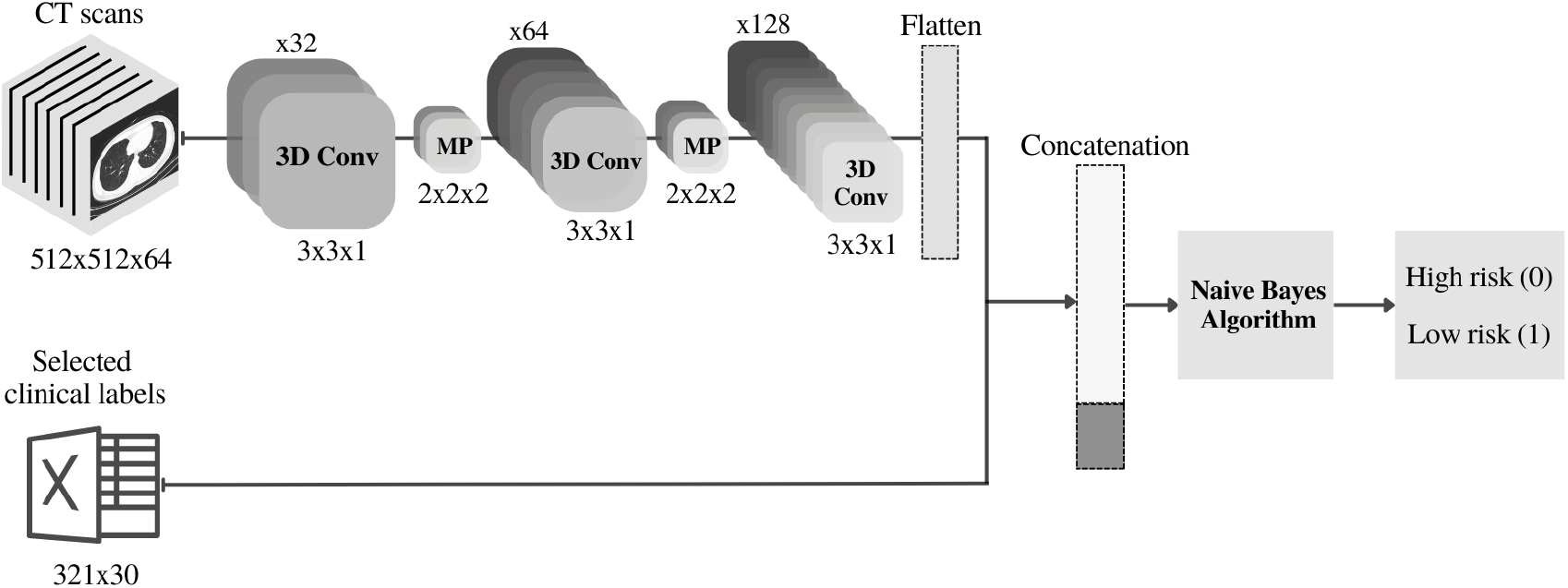
Schematic of network architucure for the terminal 3D-CNN fusion model (on CTs+30 clinical labels).

In the medial 3D-CNN approach, the extracted features-vectors of CT scans using the 3D-CNN in the previous structure were medially combined with extracted features from the original clinical data (67 clinical labels from each patient) using a 1D-CNN to create a more comprehensive features-set (Figure 4). Two fully-connected layers were finally used at the end of this structure and the output was fed into a final classification layer.

**Figure 4.**
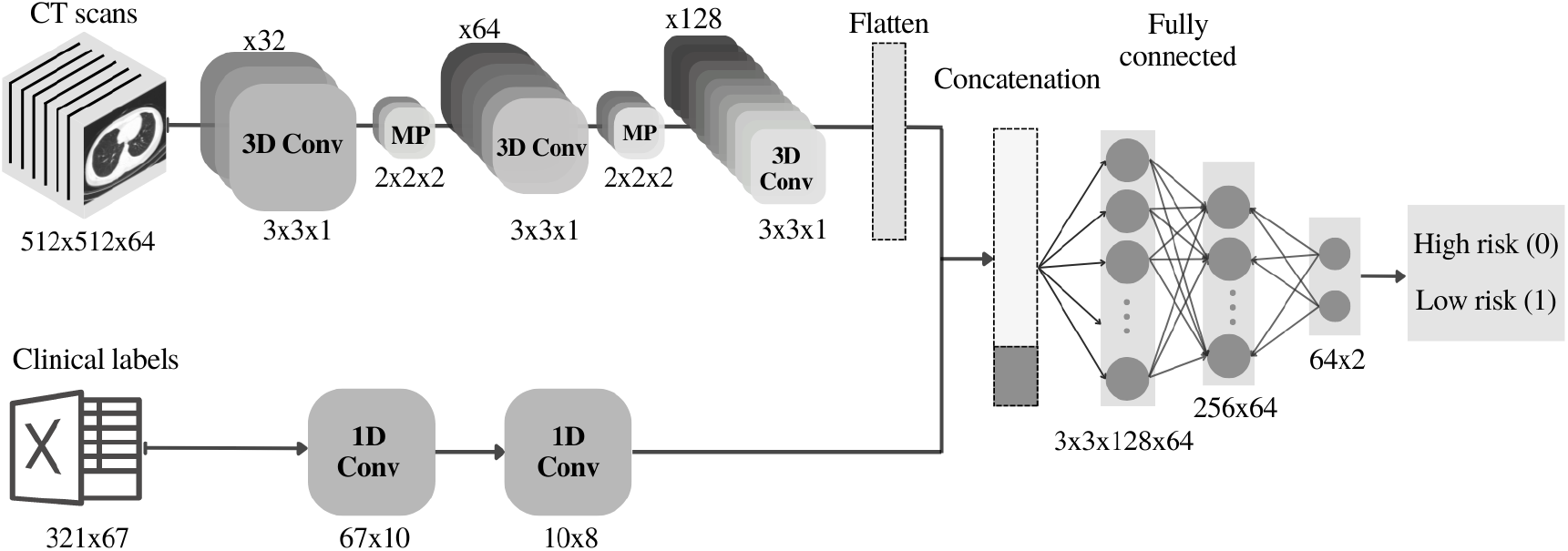
Schematic of network architucure for the medial 3D-CNN fusion model (on CTs+67 clinical labels).

##### 3.3.3.2 3D Swin Transformer models on fusion data

The complementary 3D fusion data from section 3.2.3 were used as inputs (512×512x(64+N) frames) to the 3D Video Swin Transformer to assess effectivity of data fusion approach. The schematic of our proposed data-fusion-based approach fed into the 3D Swin Transformer models is shown in Figure 5. We tested the performance of the 3D Swin Transformer model under two different scenarios for N=30 (associated with the selected clinical labels in section 3.2.1.3) and N=67 (associated with all clinical labels).

**Figure 5.**
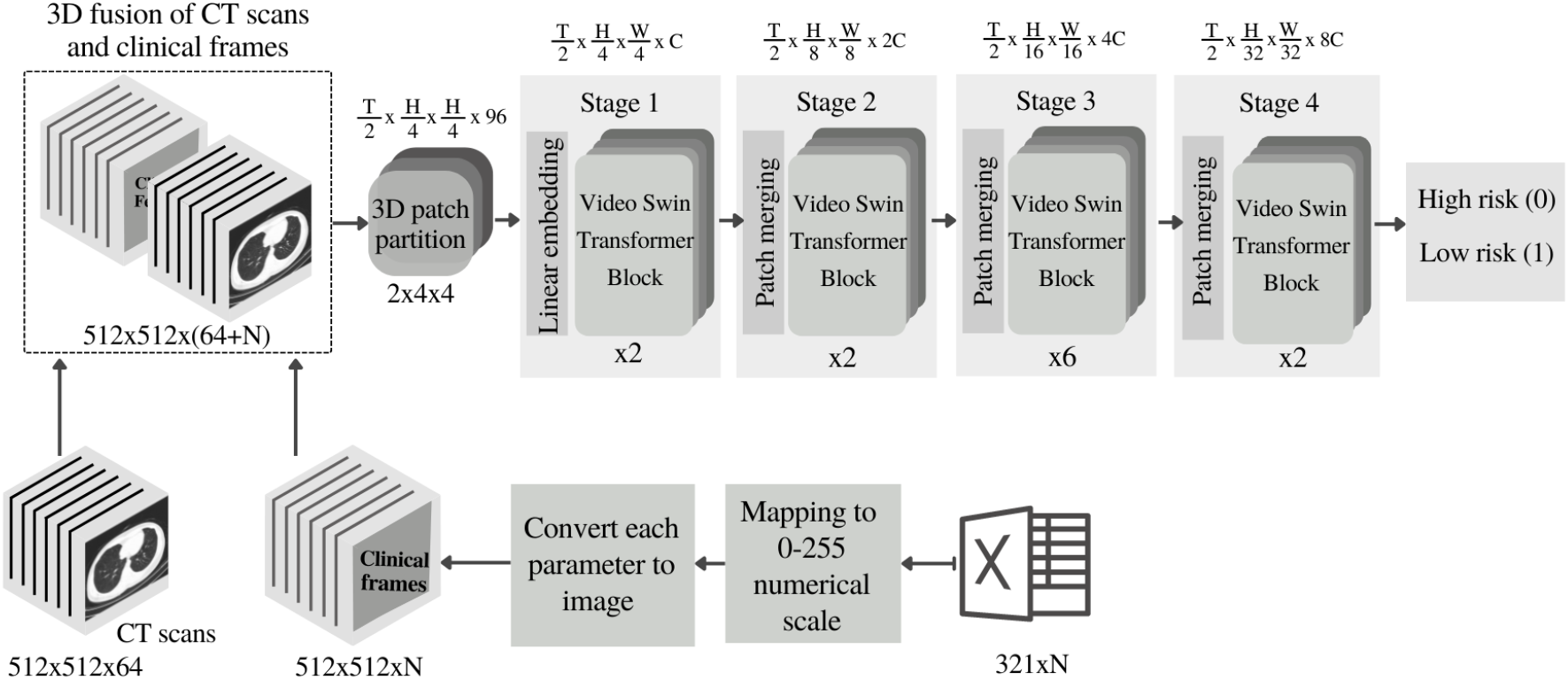
Schematic of the 3D Swin Transformer model fed with the fusion of CT scan images and clinical data. “N” denotes the number of cliniacl labels.

## 4. Performance measure

### 4.1. Performance measure selection and assessments

A k-fold cross-validation approach (k=5) was used for overall performance assessments of all models. We also used the StratifiedKFold, a stratified cross-validator algorithm, to split the imbalanced dataset into train/test sets across 5 folds.

Performance measures such as Kappa and F0.5 score could provide better validation evaluations for the classification of imbalanced datasets compared to the standard conventional measures such as “accuracy” and/or “precision/recall”. In fact, the later measures may not be reliable criteria when classification is performed on un-balanced data or when data is not normally distributed [62, 63]. AUC measure, however, includes the proportional impacts of the precision and recall metrics in validation assessments. Also, false positive rate (FPR) is clinically a critical measure (e.g., compared to true positive rate (TPR)); this is mainly because this measure indicates how many of high-risk labels have been incorrectly classified in the low-risk class. Clinically, a high FPR rate is not acceptable as the misidentification of high-risk labels can be dangerous for patients who require treatments. Due to these reasons, our performance evaluation policy was focused on models that simultaneously achieved a minimal FPR, a higher TPR, a higher AUC, and higher F0.5 score and Kappa. This “trade-off” strategy was mainly targeted to find a model with the lowest missed/wrong identifications for the high-risk class. These performance measures are described in the following.

#### 4.1.1 Kappa

Kappa statistic is a performance measure that penalizes all positive or all negative predictions in its scoring regime. This approach is especially useful in multi-class imbalanced data classification and has been therefore commonly used in datasets with imbalanced classes [64, 65]. Moreover, Kappa has been shown to provide better insights than other metrics on detecting performance variations due to drifts in the distributions of the data classes. Kappa statistic ranges between -100 (total disagreement) through 0 (default probabilistic classification) to 100 (total agreement):

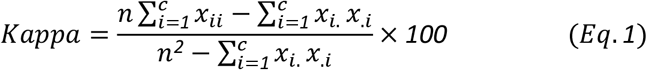

where *x*_*ii*_ is the count of cases in the main diagonal of the confusion matrix (successful predictions), *n* is the number of examples, c is the number of classes, and *x*_.*i*_, *x*_*i*._ are the column and row total counts, respectively.

#### 4.1.2 TPR, FPR, and Precision

The TPR (also called sensitivity or recall), in this article, indicates how many of the data are correctly classified in the low-risk group (class 1) while the FPR indicates how many of the data in the high-risk group are incorrectly classified in the low-risk group (class 1). Also, precision (or positive predictive value (PPV)) evaluates the number of TPs out of the total number of positive predictions which indicates how good the model was able to make positive predictions.

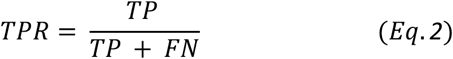

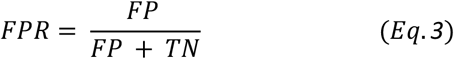

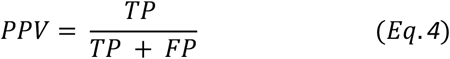

#### 4.1.3 F0.5 score

F0.5 score is the weighted version of F1 score where more weight is considered to precision than to recall (Equation 5). This is particularly important where more weight needs to be assigned to PPV for situations where FPs are considered worse than FNs.

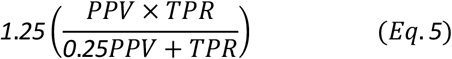

## 5. Computing infrastructure

We used New Zealand eScience Infrastructure (NeSI) high-performance computing facilities’ Cray CS400 cluster for training and testing the models. The training process was executed using enhanced NVIDIA Tesla A100 PCIe GPUs with 40 GB HBM2 stacked memory bandwidth at 1555 GB/s. Intel Xeon Broadwell CPUs (E5-2695v4, 2.1 GHz) were used on the cluster for handling the GPU jobs. The algorithms were run under Python environments (Python 3.7) using Pytorch deep learning framework (Pytorch 1.11).

## 6. Results

This section provides the obtained results for the pre-processing, clinical-data-only trained models, CT scans-only trained models, as well as the fusion approaches for both CNNs and Transformer models.

### 6.1 Pre-processed clinical data

Full results from the feature selection and feature extraction in section 3.2.1.3 using the seven conventional classification algorithms for the 1) 67 original clinical labels, 2) 13 selected clinical labels from SelectKBest algorithm, 3) 30 selected clinical labels from ExtraTree classifier, and 4) 25 extracted features from PCA algorithm are shown in Tables D.1 to D.4 in Appendix D, respectively. A trade-off performance criterion for a lower FPR and a higher TPR, F0.5 score, and Kappa in these tables showed that the Gaussian Naïve bays (NB) performed much better across the four approaches above. The abstracted results in Table 1 further confirm that the classification of the clinical data using Gaussian NB fed with 30 selected clinical labels from the ExtraTree classifier has led to better performances compared to other approaches.

**Table 1.**
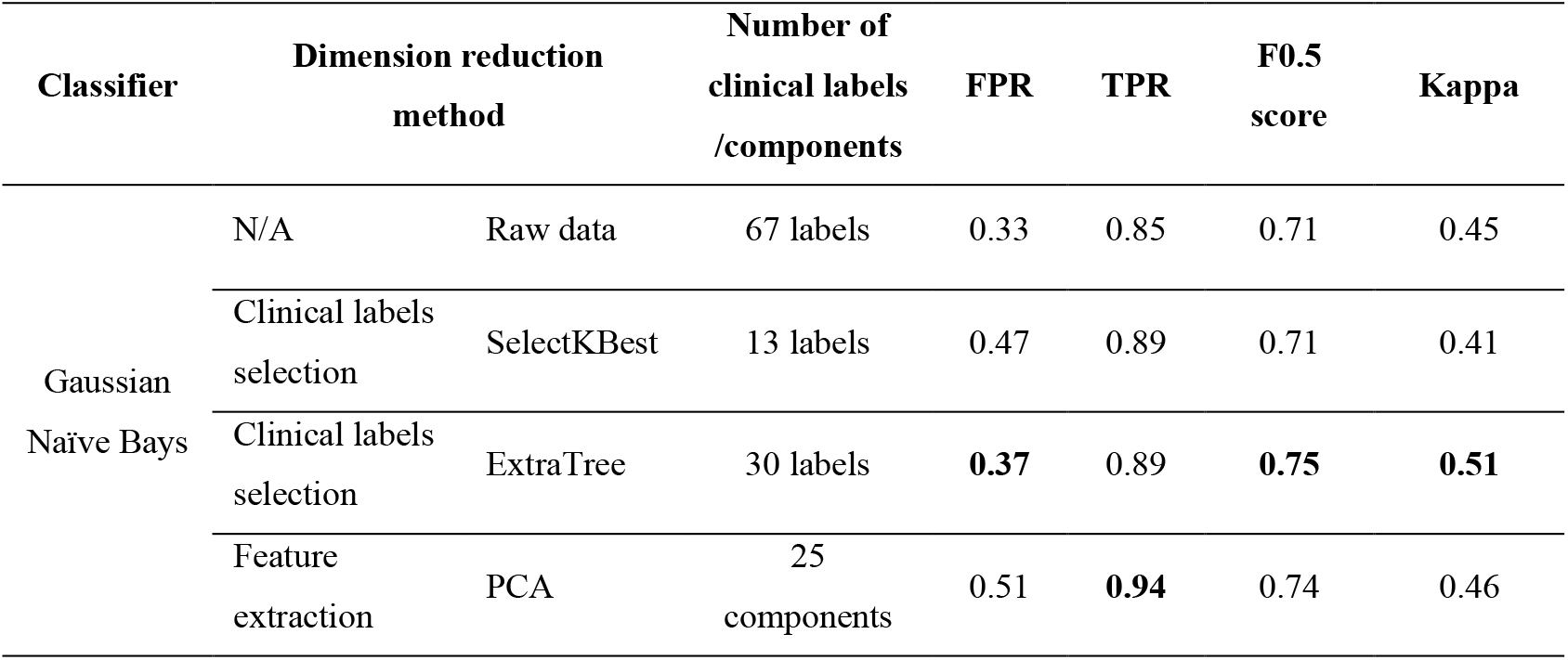
Comparison between the dimension reduction methods on the clinical data

In addition, features-space assessments using the t-SNE algorithm on the four above schemes are shown in Figure 6A to 6D, respectively. The features-space plots in this figure hold high-complexity and a visual binary classification seems to be a challenging due to the negligible differences between the images. Nevertheless, the application of t-SNE on the 30 selected labels from ExtraTree classifier in Figure 6C seems to provide a much better visually classifiable data. Due to the above reasons, the dataset containing the 30 selected clinical labels from ExtraTree classifier were used as the clinical dataset for models in section 3.3.1 and 3.3.3, where this data were further fused with the CT images to shape the fusion datasets.

**Figure 6.**
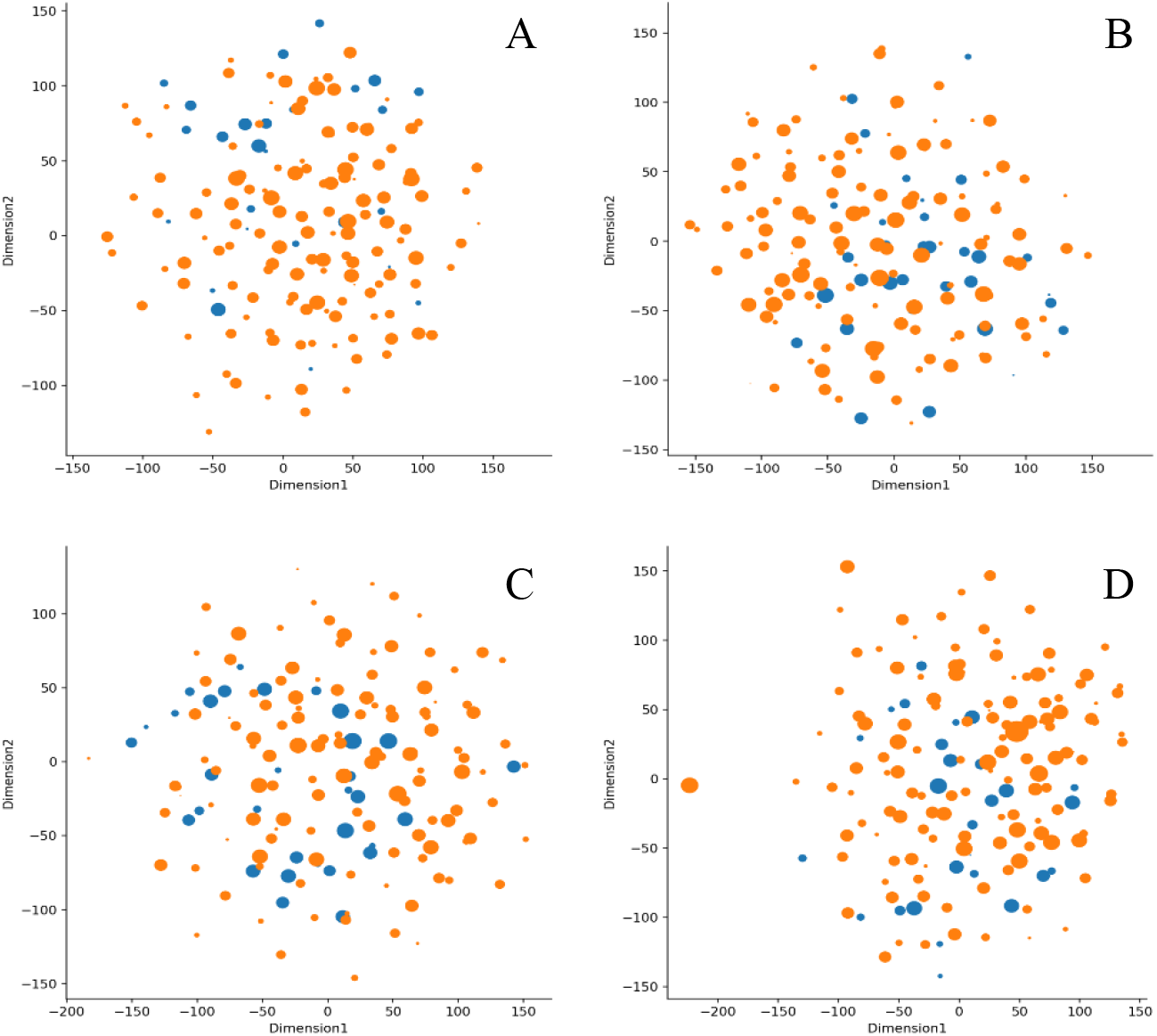
Visual representations from the t-SNE approach using A: the main dataset including all 67 clinical labels (with no dimension reduction), B: 13 selected clinical labels from SelectKBest, C: 30 selected clinical labels from ExtraTree classifier, and D: 25 extracted features from PCA. blue: high-risk (class 0), orange: low-risk (class 1).

### 6.2 Models on the clinical data only

Results from the seven classification algorithms in 3.3.1 are shown in Table 2. Each classifier was assessed using the 30 selected clinical labels from ExtraTree classifier. As shown, here the gradient boosting algorithm has outperformed the other algorithms.

**Table 2.** Performance results of the classifiers

| Model category | Model name | FPR | TPR | F0.5 score | ROC/AUC* | Recall | Precision | Kappa (-1, 1) |
| --- | --- | --- | --- | --- | --- | --- | --- | --- |
| Section 3.3.1<br>**Clinical data only**<br>(on the set of 30 selected clinical labels from ExtraTree classifier) | Gaussian NB | 0.49 | 0.70 | 0.57 | 0.60 | 0.61 | 0.59 | 0.19 |
|  | Random Forest | 0.07 | 0.56 | 0.60 | 0.74 | 0.74 | 0.64 | 0.27 |
|  | Gradient Boosting | 0.14 | 0.70 | 0.66 | 0.78 | 0.78 | 0.68 | 0.37 |
|  | XGBRF | 0.16 | 0.65 | 0.62 | 0.72 | 0.73 | 0.64 | 0.28 |
|  | k-nearest neighbors | 0.47 | 0.58 | 0.50 | 0.55 | 0.56 | 0.52 | 0.05 |
|  | SVM | 0.18 | 0.18 | 0.32 | 0.50 | 0.50 | 0.42 | 0 |
|  | MLP | 0.40 | 0.40 | 0.22 | 0.50 | 0.50 | 0.20 | 0 |
| Section 3.3.2<br>**CTs only** | 3D-CNN | 0.83 | 0.84 | 0.63 | 0.57 | 0.57 | 0.78 | 0.22 |
|  | 3D Swin Transformer | 0.38 | 0.89 | 0.75 | 0.75 | 0.75 | 0.75 | 0.49 |
| Section 3.3.3<br>**Data fusion** | Terminal 3D-CNN on CTs+30 labels | 0.36 | 0.90 | 0.75 | 0.76 | 0.76 | 0.76 | 0.51 |
|  | Medial 3D-CNN on CTs+67 labels | 0.65 | 0.98 | 0.70 | 0.66 | 0.66 | 0.67 | 0.37 |
|  | 3D Swin Transformer on CTs+30 labels | 0.35 | 0.91 | 0.78 | 0.78 | 0.78 | 0.80 | 0.55 |
|  | 3D Swin Transformer on CTs+67 labels | 0.40 | 0.95 | 0.82 | 0.77 | 0.77 | 0.83 | 0.60 |

\***ROC**: Receiver Operating Characteristics Curve, **AUC**: Area under the ROC Curve

### 6.3 Models trained on CT images only

Results of the 5-fold cross-validation from the 3D-CNN and 3D Swin Transformer models in section 3.3.2 (trained on the CT-images only) are shown in Table 2. Results from the Transformer model on the CT images only shows improvement for all measures including FPR (0.45 lower), Kappa (0.27 higher), and F0.5 score (0.11 higher) compared to the 3D-CNN.

### 6.4 Models trained on fusion data

Results of the 5-fold cross-validation for each of the data-fusion approaches in sections 3.3.3 are also shown in Table 2. As shown, the Transformer fusion models as well as the Terminal 3D-CNN have resulted in improved overall scores, across all measures, compared to the medial 3D-CNN fusion approach.

ROC curves of the top performing models from each section as well as the top performing 3D-CNN on the fusion data are shown in Figure 7.

**Figure 7.**
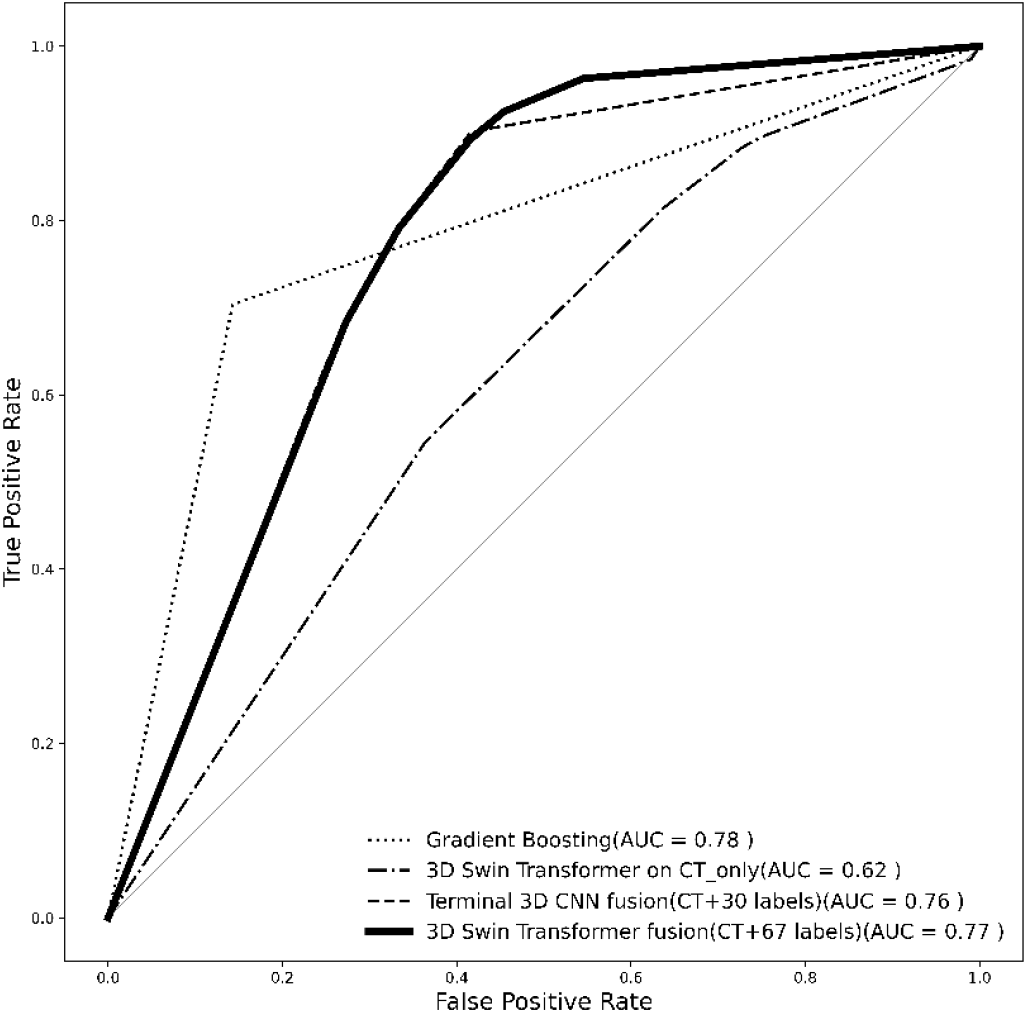
Mean ROC curves from the top performing models. Bold line: 3D Swin Transformer on fusion data (CT+67 labels), dashed-dotted line: 3D Swin Transformer on CTs only, dotted line: Gradient Boosting, dashed line: Terminal 3D-CNN fusion (CT+30 labels).

## 7. Discussion and conclusion

This paper, for the first time, demonstrated how a 3D data fusion approach of combining CT scan images and patients’ clinical data can help to improve the performance of Visual Transformer and CNN models for predicting high-risk Covid-19 infection. Other studies have mainly focused on feeding such networks with either CT scan images or patients’ clinical data. The paper explored a comprehensive set of strategies to evaluate optimal predictive model across a number of classifiers tested on a relatively large dataset of 380 patients. This research demonstrates the superiority of data-fusion approaches used in 3D Swin Transformers for better identification of high-risk Covid-19 infected patients.

Here we showed that the performance of a 3D Swin Transformer model tested on the fusion of CT scan images and the original set of 67 clinical labels outperformed all other strategies in this work (FPR=0.40, TPR=0.95, F0.5 score = 0.82, AUC=0.77, Kappa=0.60) where the models were fed with fusion-type datasets, CT scan images only, and clinical data only. Here, we formed 3D fusion datasets by re-shaping the clinical data into 512×512x’number of clinical labels’ format and combined them with CT scan images of size 512×512×64 to create our fusion dataset. It is inferred that our strategy for the dimension expansion of clinical data and fusing them with the CT scan images has successfully helped the self-attention layers within the Swin Transformer model to effectively rate interconnectivity between the clinical data and the CT images for better classifications.

We further tested and compared the performance of a terminal 3D-CNN model (on the CT+30 clinical labels), a medial 3D-CNN (on the CT+67 clinical labels), 3D Swin Transformers (on the CT+30 clinical labels and on the CT+67 clinical labels, respectively) to the original approach. Results from Table 2 indicates that our selected set of 30 clinical labels from the original pool of 67 clinical labels fused with the patients’ CT scan images has been consistently and effectively helpful to achieve competitive performances compared to the 3D Swin Transformer on the CT+67 clinical labels. Here, the 3D Swin Transformer model on the fusion of CT+30 clinical labels achieved FPR=0.35, TPR=0.91, F0.5 score = 0.78, AUC=0.78, Kappa=0.55, and the terminal 3D-CNN model on the fusion of CT+30 clinical labels achieved FPR=0.36, TPR=0.90, F0.5 score = 0.75, AUC=0.76, Kappa=0.51. These closer performance measures from the selected set of 30 clinical labels suggest that these dominant labels may hold clinical values in the clinical settings for a better identification of the illness and could be looked at in details in future studies. These clinical labels have been listed in Table A.1 of appendix A. We also assessed classification capabilities of a 3D-CNN model and a Video Swin Transformer on sets of 3D CT scan images only. Here, the 3D Swin Transformer achieved much better results (FPR=0.38, TPR=0.89, F0.5 score = 0.75, AUC=0.75, Kappa=0.49) compared to the 3D-CNN with higher FPR, lower AUC and Kappa (FPR=0.83, TPR=0.84, F0.5 score = 0.63, AUC=0.57, Kappa=0.22).

Our assessments also showed that conventional classifiers, fed with patients’ clinical data only, poorly classified the data compared to the other two approaches above, namely the fusion and CT-only strategies (see Table 2). Amongst the conventional classifiers, Gradient boosting was found to outperform the other ones when only fed with the clinical data (FPR=0.14, TPR=0.70, F0.5 score = 0.66, AUC=0.78, Kappa=0.37).

An overall trade-off assessment shows that the 3D Swin Transformer fed with the fusion of CT scan images and the full set of 67 clinical labels identified high-risk patients from the low-risk class more accurately compared to the other approaches. This was closely followed by 3D Swin Transformer model fed with a fusion of CT images and the set of 30 selected clinical labels from ExtraTree classifier. The 3D Swin Transformer again demonstrated superiority compared to the 3D-CNN approach, even when both models were fed with CT images only; however, the overall performance was found to be lower than the data-fusion approach. Classification performances remarkably decreased across all the seven conventional models when only the clinical data were used. The mean ROC curves in Figure 6 from the top performing models in each section demonstrate how the 3D Swin Transformer on fusion data (CT+67 labels) outperformed the other approaches. We have also provided the ROC curve of the top performing 3D-CNN model on the fusion data, namely the Terminal 3D-CNN fusion (CT+30 labels) to show how the choice of 30 selected clinical labels could also help our proposed CNN-based model to achieve competitive performances compared to the 3D Swin Transformer on the fusion data.

Overall, we expect potential clinical utility for the proposed 3D Video Swin Transformer fed with fusion datasets from patients’ CT images and clinical data for reliable prediction of outcomes in Covid-19-infected patients. The improved performances of the Transformer models show robust capability for a future validation study on larger datasets. We encourage readers to apply the proposed fusion scheme in this work to larger clinical datasets for further validity assessments. Results from this research highlight the possibilities of predicting the severity of Covid-19 infection, at the time of admission to the clinical centers, when effectivity of early treatments is evident.

## Conclusion

This paper demonstrated how the performance of Visual Transformers, namely a 3D Swin Transformer, could remarkably improve for predicting Covid-19 outcomes when fed with a novel 3D data fusion approach of integrating CT scan images with patients’ clinical data. The paper further explored and compared capabilities of a series of models including Transformers (on CT images only), 3D-CNNs (both on the fusion dataset and on CT images only) as well as conventional classifiers (on the clinical data only). Results showed that the use of fusion dataset provided opportunity for the 3D Swin Transformer model to better aggregate globally and locally interconnected features of the data and perform better compared to all other models. Results confirmed that this was valid for the larger fusion dataset of 64 CT scans + 67 clinical labels and the 64 CT scans + 30 selected clinical labels. The paper further discussed how genetic algorithm (GA) is a suitable choice for hyper-parameter tuning of the 3D-CNN models. We also investigated a series of strategies to find and select a proper set of clinical labels from the pool of clinical data for the classification of imbalance data. The paper further discusses imputation techniques to deal with missing values in the dataset. Overall, this paper demonstrates possibilities of predicting the severity of outcome in Covid-19 infected individuals at or around the time of admission to hospital using fusion datasets from patients’ CT images and clinical data.

## Data Availability

The data generated and/or analyzed during the current study are not publicly available for legal/ethical reasons but are available from the corresponding author on reasonable request.

## Supporting Information

Supplementary material can be found at the end of this paper. Supporting Information is available from the authors.

## Author contribution

The algorithm development, data analysis and manuscript writing/preparation were undertaken by S.S, M.Z, and H.A. Data was collected and pre-processed by P.A, M.R, S.A.S, A.S, and S.A. Z.G contributed to algorithm development. R.A, M.G, and N.N provided technical advice. Manuscript was reviewed, edited, and revised by H.A. H.A was the main supervisor of the work and assisted with study design, administration, and implementation. The final submitted article has been revised and approved by all authors.

## Acknowledgements

- We would also like to acknowledge the use of New Zealand eScience Infrastructure (NeSI) high performance computing facilities to the results of this research. URL: https://www.nesi.org.nz.
- We would also like to thank Mohammad Arbabpour Bidgoli from the KTH Royal Institute of Technology in Sweden for assisting with data collection.

## Supplementary material

## Appendix A

**Table A.1.** Patients’ clinical measurements (mean±std). X(Y%): X represents the label counts in the population, and the associated % of the label in the population is shown with Y. *Italic labels* represent the excluded labels previouly explained in section 3.2.1.1.

| Clinical variable/label | Total (n=380) |
| --- | --- |
| **Gender** |  |
| Female | 133(35%) |
| Male | 247(65%) |
| **Age** |  |
| Mean ± SD | 53.82±17.92 |
| **Exposure history** |  |
| EHF * | 57(15%) |
| HOT * | 29(7.63%) |
| ESP * | 83(21.84%) |
| CIH * | 50(13.16%) |
| **Comorbid disease** |  |
| Diabetes Mellitus | 77(20.26%) |
| Vascular disease | 59(15.53%) |
| Obesity | 49(12.89%) |
| Smoking | 24(6.32%) |
| Kidney | 19(5.00%) |
| Asthma | 13(3.42%) |
| Other lung diseases | 23(6.05%) |
| Malignancy | 12(3.16%) |
| Hematic | 10(2.63%) |
| Rheumatology | 10(2.63%) |
| Neurological | 14(3.68%) |
| Hypertension | 47(12.37%) |
| UCC* | 12(3.16%) |
| **Clinical manifestations** |  |
| Fever | 225(59.21%) |
| Cough | 234(61.58%) |
| Dyspnea | 196(51.58%) |
| Myalgia | 179(47.11%) |
| Headache | 128(33.68%) |
| Chest pain | 81(21.32%) |
| Nausea | 101(26.58%) |
| Sputum | 83(21.84%) |
| Chills | 174(45.79%) |
| Hemoptysis | 13(3.42%) |
| Sore Throat | 15(3.95%) |
| Anorexia | 198(52.11%) |
| Rhinorrhea | 47(12.37%) |
| Anosmia | 54(14.21%) |
| Weakness | 207(54.47%) |
| Loss of consciousness | 22(5.79%) |
| Diarrhea | 70(18.42%) |
| Earache | 13(3.42%) |
| Wheezing | 0(0%) |
| Arthralgia | 31(8.16%) |
| Respiratory distress | 5(1.32%) |
| Dizziness | 63(16.58%) |
| Convulsion | 6(1.58%) |
| Abdominal pain | 32(8.42%) |
| Conjunctivitis | 23(6.05%) |
| Rash | 3(0.79%) |
| Skin lesion | 1(0.26%) |
| Lymphadenopathy | 0(0%) |
| Sweating | 19(5.00%) |
| Hematochezia | 2(0.53%) |
| Cold sweating | 14(3.68%) |
| **Venous blood gas analysis** |  |
| WBC (10<sup>9</sup>/L) | 7.19±5.18 |
| Hemoglobin (g/dl) | 13.04±1.20 |
| Hematocrit (%) | 38.87±5.14 |
| Platelet (10<sup>9</sup>/L) | 194.42±83.32 |
| Lymphocyte (%) | 21.39±11.75 |
| Neutrophil (%) | 71.63±12.99 |
| *Lactate Dehydrogenase (LDH)* | 640.125±382.65 |
| **Complete blood count** |  |
| pH | 7.41±0.1 |
| PO<sub>2</sub> (mm Hg) | 35.19±19.42 |
| PCO<sub>2</sub> (mm Hg) | 41.62±10.33 |
| HCO<sub>3</sub> (mEq/L) | 26.31±7.10 |
| O<sub>2</sub>satVBG | 58.87±22.15 |
| **Kidney enzymes** |  |
| Urea (mg/dL) | 47.40±38.33 |
| Creatinine (mg/dL) | 1.46±1.33 |
| **Others** |  |
| Sodium (mEq/L) | 136.90±7.75 |
| CRP (mg/L) * | 53.00±48.54 |
| *Potassium (mEq/L)* | 4.06±0.68 |
| Calcium (mg/dL) | 8.27±1.08 |
| *Magnesium (mg/dL)* | 3.01±9.35 |
| *ESR (mm/hr) \** | 49.42±27.28 |
| *CPK (U/L) \** | 317.18±743.51 |
| *Blood sugar(mg/dL)* | 153.97±70.74 |
| *Bill Total* | 7.07±38.02 |
| *Procalcitonin (ng/ml)* | 1.05±1.88 |
| *PCR \** | 94(92.16%) |
| **Presenting vital sign** |  |
| *Temperature (c)* | 36.91±3.31 |
| *Systolic BP (mmHg)* | 86.46±32.69 |
| *Diastolic BP (mmHg)* | 106.90±25.06 |
| *Respiratory rate (/min)* | 18.69±4.46 |
| *Heart rate (/min)* | 90.05±18.00 |
| *Saturation O<sub>2</sub> (%)* | 92.15±7.67 |
| **Coagulation profile** |  |
| *PT (s)\** | 12.99±3.12 |
| *PTT (s)\** | 28.85±11.86 |
| *INR (IU)\** | 2.01±8.86 |
| **Liver enzymes** |  |
| *AST (U/L)\** | 66.52±204.179 |
| *ALT (U/L)\** | 38.95±35.60 |

\*Exposure to healthcare facilities (EHF) history of traveling (HOT) exposure to the suspected patient (ESP) covid-19 infection in household (CIH) Use of corticosteroid for comorbidities (UCC) c-reactive protein (CRP) erythrocyte sedimentation rate (ESR) Creatine phosphokinase (CPK) polymerase chain reaction (PCR) Prothrombin time (PT) Partial thromboplastin time (PTT) international normalized ratio (INR) aspartate aminotransferase (AST) alanine transaminase (ALT)

## Appendix B

**Figure B.1.**
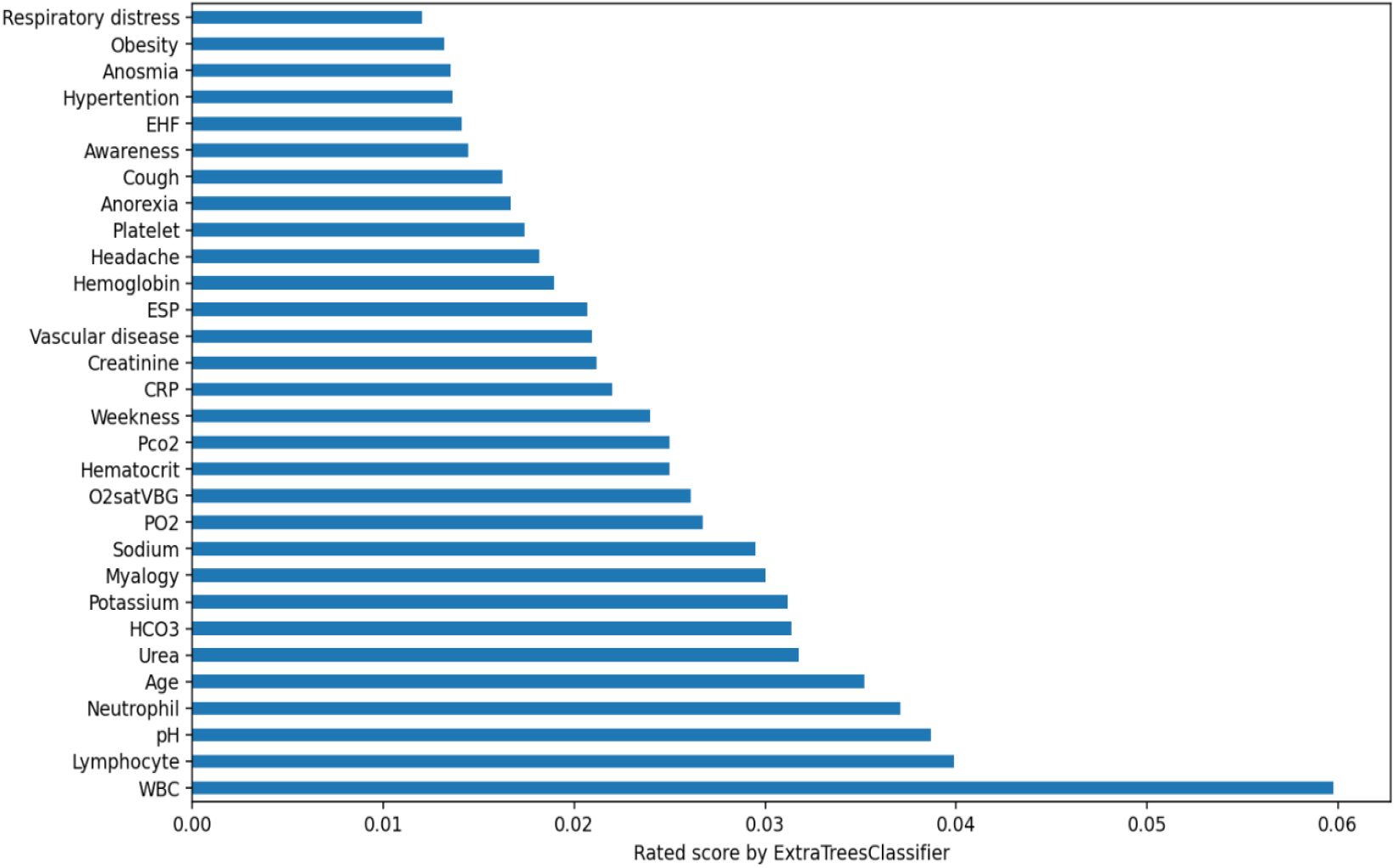
The suggested set of 30 selected clinical labels from ExtraTree classifier.

**Figure B.2.**
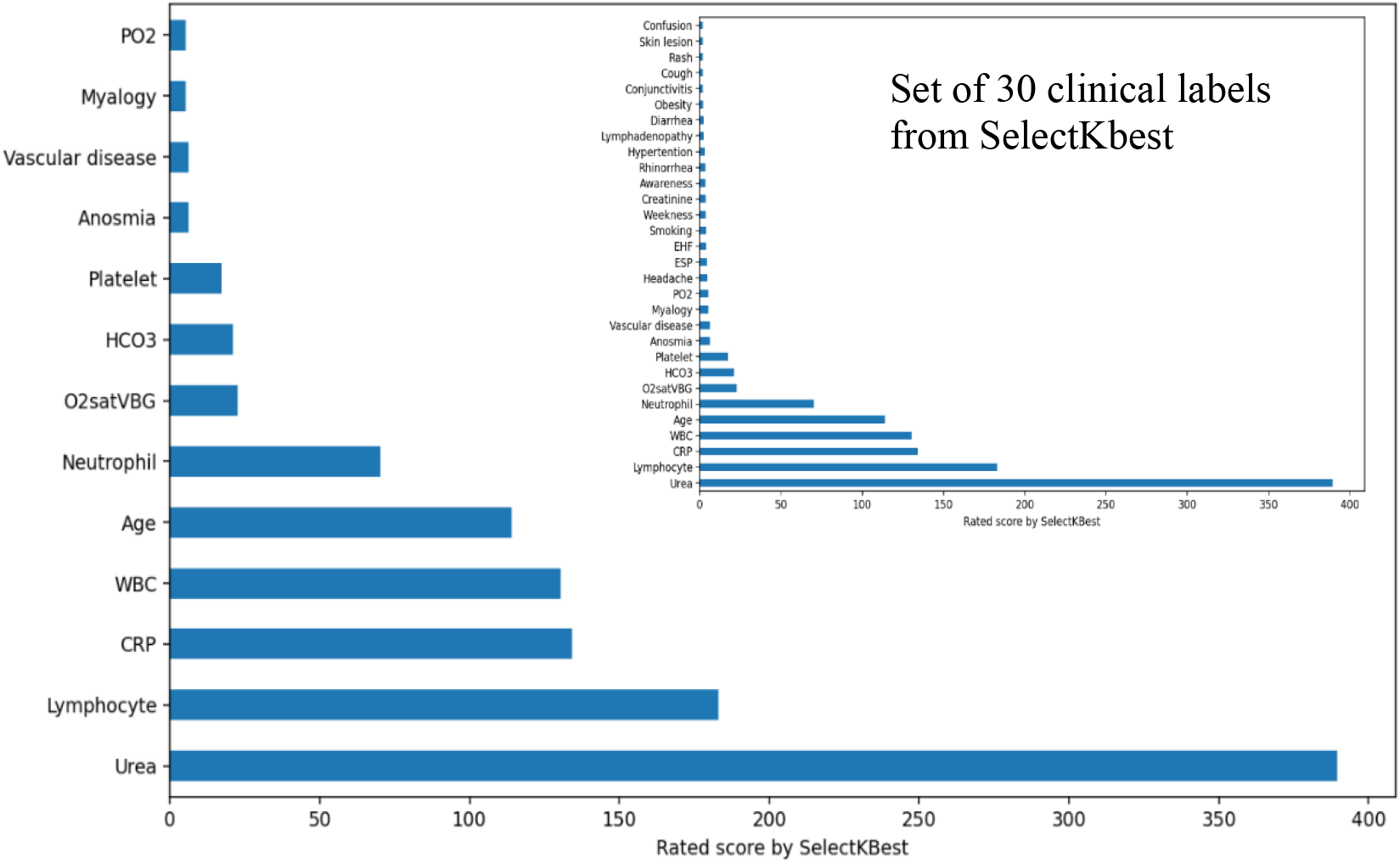
The suggested set of 13 clinical labels from SelectKbest algorithm.

## Appendix C

**Table C.1.**
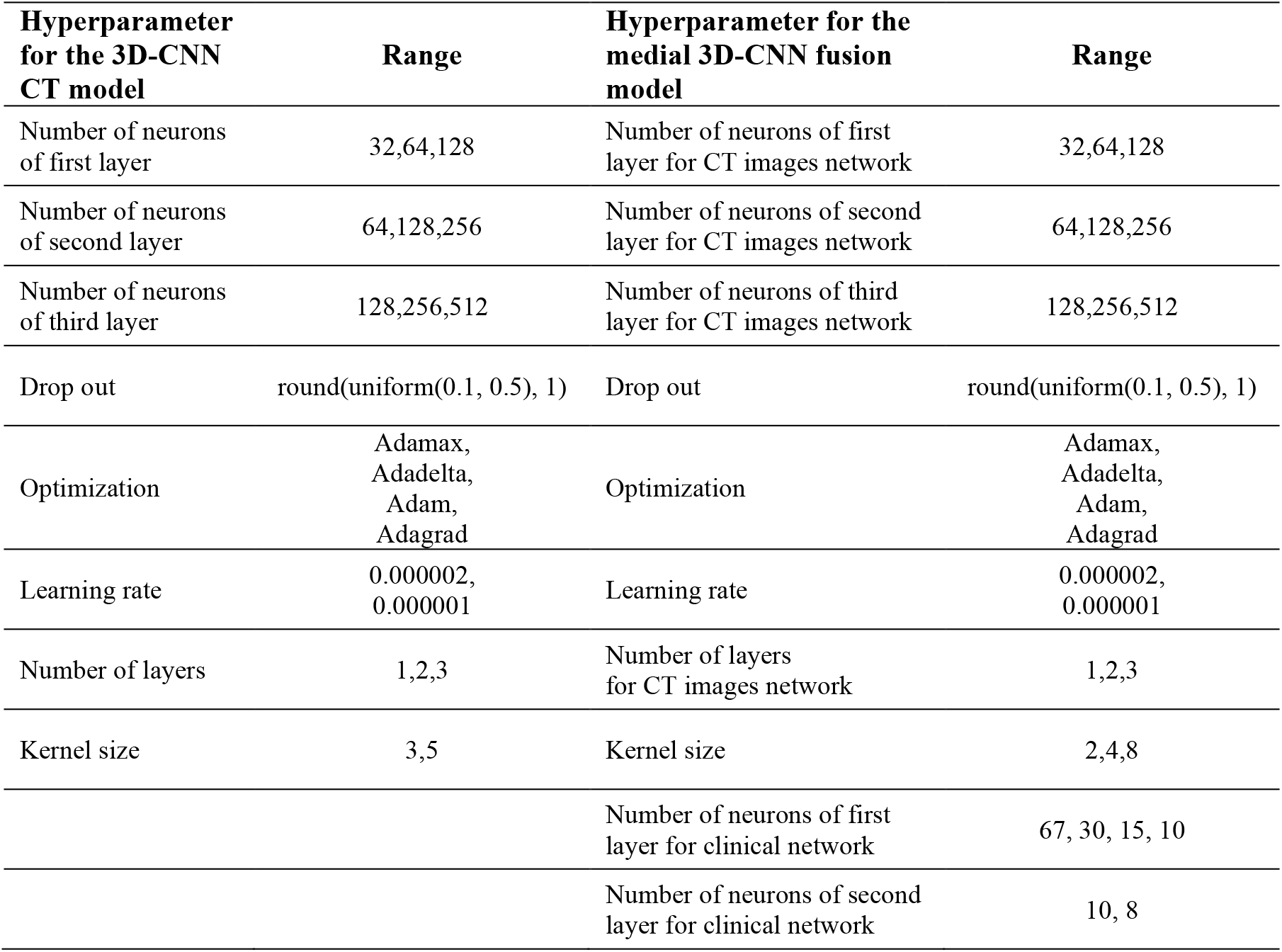
Selected hyperparameters from the Genetic Algorithm for the 3D-CNN CT model (section 3.3.2.1) and the medial 3D-CNN fusion model (section 3.3.3.1).

| **Hyperparameter for the 3D-CNN CT model** | **Range** | **Hyperparameter for the medial 3D-CNN fusion model** | **Range** |
| --- | --- | --- | --- |
| Number of neurons of first layer | 32,64,128 | Number of neurons of first layer for CT images network | 32,64,128 |
| Number of neurons of second layer | 64,128,256 | Number of neurons of second layer for CT images network | 64,128,256 |
| Number of neurons of third layer | 128,256,512 | Number of neurons of third layer for CT images network | 128,256,512 |
| Drop out | round(uniform(0.1, 0.5), 1) | Drop out | round(uniform(0.1, 0.5), 1) |
| Optimization | Adamax, Adadelata, Adam, Adagrad | Optimization | Adamax, Adadelata, Adam, Adagrad |
| Learning rate | 0.000002, 0.000001 | Learning rate | 0.000002, 0.000001 |
| Number of layers | 1,2,3 | Number of layers for CT images network | 1,2,3 |
| Kernel size | 3,5 | Kernel size | 2,4,8 |
|  |  | Number of neurons of first layer for clinical network | 67, 30, 15, 10 |
|  |  | Number of neurons of second layer for clinical network | 10, 8 |

## Appendix D

**Table D.1.** Results of the seven conventional algorithms in section 3.2.1 on the 67 original clinical labels.

| Classifier | FPR | TPR | Recall | Precision | F0.5 score | Kappa | Training accuracy | Test accuracy |
| --- | --- | --- | --- | --- | --- | --- | --- | --- |
| SVM | 1 | 1 | 0.5 | 0.41 | 0.43 | 0 | 0.82 | 0.82 |
| MLP | 0.62 | 0.95 | 0.67 | 0.76 | 0.73 | 0.4 | 0.99 | 0.85 |
| KNN | 0.56 | 0.81 | 0.63 | 0.62 | 0.61 | 0.23 | 0.92 | 0.75 |
| Gaussian NB | 0.33 | 0.85 | 0.76 | 0.71 | 0.71 | 0.45 | 0.83 | 0.82 |
| XGBoost | 0.6 | 0.95 | 0.68 | 0.78 | 0.73 | 0.41 | 0.89 | 0.89 |
| Random Forest | 0.65 | 0.97 | 0.66 | 0.79 | 0.73 | 0.39 | 1 | 0.86 |
| Gradient Boosting | 0.54 | 0.92 | 0.69 | 0.72 | 0.71 | 0.39 | 0.97 | 0.97 |
| AVG | 0.61 | 0.92 | 0.65 | 0.68 | 0.66 | 0.32 | 0.92 | 0.85 |

**Table D.2.** Results of the seven conventional algorithms in section 3.2.1 on the 13 selected clinical labels from SelectKBest algorithm.

| Classifier | FPR | TPR | Recall | Precision | F0.5 score | Kappa | Training accuracy | Test accuracy |
| --- | --- | --- | --- | --- | --- | --- | --- | --- |
| SVM | 1 | 1 | 0.5 | 0.41 | 0.43 | 0 | 0.82 | 0.82 |
| MLP | 0.57 | 0.92 | 0.68 | 0.71 | 0.7 | 0.37 | 0.94 | 0.83 |
| KNN | 0.58 | 0.81 | 0.62 | 0.61 | 0.6 | 0.21 | 0.92 | 0.74 |
| Gaussian NB | 0.47 | 0.89 | 0.71 | 0.71 | 0.71 | 0.41 | 0.85 | 0.83 |
| XGBoost | 0.6 | 0.96 | 0.68 | 0.81 | 0.75 | 0.42 | 1 | 0.86 |
| Random Forest | 0.67 | 0.96 | 0.65 | 0.81 | 0.71 | 0.35 | 1 | 0.85 |
| Gradient Boosting | 0.58 | 0.94 | 0.68 | 0.75 | 0.72 | 0.4 | 0.98 | 0.98 |
| AVG | 0.64 | 0.92 | 0.64 | 0.69 | 0.66 | 0.31 | 0.93 | 0.84 |

**Table D.3.** Results of the seven conventional algorithms in section 3.2.1 on the 30 selected clinical labels from ExtraTree classifier.

| Classifier | FPR | TPR | Recall | Precision | F0.5 score | Kappa | Training accuracy | Test accuracy |
| --- | --- | --- | --- | --- | --- | --- | --- | --- |
| SVM | 1 | 1 | 0.5 | 0.41 | 0.43 | 0 | 0.82 | 0.82 |
| MLP | 0.53 | 0.92 | 0.7 | 0.75 | 0.72 | 0.42 | 0.94 | 0.84 |
| KNN | 0.54 | 0.81 | 0.64 | 0.63 | 0.62 | 0.24 | 0.92 | 0.75 |
| Gaussian NB | 0.37 | 0.89 | 0.76 | 0.75 | 0.75 | 0.51 | 0.86 | 0.83 |
| XGBoost | 0.56 | 0.96 | 0.7 | 0.8 | 0.76 | 0.46 | 1 | 0.87 |
| Random Forest | 0.67 | 0.97 | 0.65 | 0.83 | 0.73 | 0.37 | 1 | 0.86 |
| Gradient Boosting | 0.58 | 0.92 | 0.67 | 0.72 | 0.7 | 0.37 | 0.97 | 0.97 |
| AVG | 0.61 | 0.92 | 0.66 | 0.7 | 0.67 | 0.34 | 0.93 | 0.85 |

**Table D.4.** Results of the seven conventional algorithms in section 3.2.1 on the 25 extracted features from PCA algorithm.

| Classifier | FPR | TPR | Recall | Precision | F0.5 score | Kappa | Training accuracy | Test accuracy |
| --- | --- | --- | --- | --- | --- | --- | --- | --- |
| SVM | 1 | 1 | 0.5 | 0.41 | 0.43 | 0 | 0.82 | 0.82 |
| MLP | 0.62 | 0.94 | 0.66 | 0.77 | 0.72 | 0.38 | 0.99 | 0.84 |
| KNN | 0.56 | 0.81 | 0.63 | 0.62 | 0.61 | 0.23 | 0.92 | 0.75 |
| Gaussian NB | 0.51 | 0.94 | 0.71 | 0.76 | 0.74 | 0.46 | 0.87 | 0.86 |
| XGBoost | 0.74 | 0.97 | 0.62 | 0.8 | 0.7 | 0.31 | 1 | 0.85 |
| Random Forest | 0.82 | 0.99 | 0.58 | 0.87 | 0.66 | 0.24 | 1 | 0.85 |
| Gradient Boosting | 0.77 | 0.95 | 0.59 | 0.69 | 0.64 | 0.22 | 1 | 0.82 |
| AVG | 0.72 | 0.94 | 0.61 | 0.7 | 0.64 | 0.26 | 0.94 | 0.83 |

